# Real-World Outcomes of a Traffic-Light Approach to Lifestyle Modification in Chronic Disease: Evidence from an Integrative Health Clinic

**DOI:** 10.64898/2026.11.10.26367386

**Authors:** Rakesh Sarwal, Kalidas Dattatraya Chavan, Jagadish C. Sharma, Rajesh Manocha

## Abstract

**Background:** Non-communicable diseases driven by unhealthy lifestyles are a growing and largely avoidable burden among industrial workers. Patient-level evidence on the effectiveness of structured lifestyle programmes delivered alongside standard care — such as a traffic-light dietary guideline, a Yoga protocol and a daily four-spice culinary infusion — remains limited. The Integrative Health Clinic (IHC) at ESIC Medical College & Hospital, Faridabad, India, is a family-medicine-style outpatient clinic that follows the 21-item Health Building Lifestyle Guideline and delivers care through a 7-step COM-B-based protocol. We therefore estimated patient-reported relief and compliance with three lifestyle practices in patients attending the IHC.

**Methods:** We conducted a retrospective single-arm cohort of patients first seen between February 2025 and 30 September 2026, with follow-up through 4 November 2026. All patients received the three-component lifestyle intervention (advice on spice infusion, diet and Yoga) and were offered follow-up; the 1,466 patients with a usable telephone contact were called for feedback and reinforcement of practices. A physical visit with a coded diagnosis was treated as a valid record. Each patient called was coded once for relief and for spice, diet and Yoga compliance using the most recent definitive answer (YES/NO); calls without a definitive answer (“no response”, “not interested”) were skipped, and patients with no definitive answer on any call were classified as such. Disease burden, including the screening-detected diagnoses, was counted as patient–condition pairs and classed into 14 systems.

**Results:** There were 4,047 valid visit records from 1,737 unique patients (904 female [52.0%]; mean age 40.3 ± 14.6 years; 833 male [48.0%]; mean age 44.8 ± 15.1 years). Of these, there were 1,035 single-visit [59.6%] and 702 revisit patients [40.4%], i.e. 2.33 visits per patient. The 1,466 analysed patients (763 female [52.1%] and 703 male [47.9%]) comprised 666 revisit and 800 single-visit patients. Their most frequent patient–condition pairs were hypertension (511), obesity (397) and arthritis (268); objective-evidence screening contributed 246 pairs in 233 patients (obesity 161, hypertension 39, anaemia 44, diabetes 2). Of the 1,466 (84.4% of 1,737) patients with a usable telephone contact who were called, 1,016 had a definitive relief answer: 850 (83.7%; 95% CI 81.3–85.8) reported relief and 166 reported no relief. The remaining 450 had no definitive answer. Among the 1,016 patients with a definitive answer, relief was 87.2% (395/453) in revisit versus 80.8% (455/563) in single-visit patients. Treating all 450 patients without a definitive relief answer as no relief, the conservative lower bound of patients reporting relief was 58.0% overall (850/1,466), 59.3% in revisit (395/666) and 56.9% in single-visit patients (455/800). Among the 1,016 patients with practice data, compliance with the spice infusion, diet and Yoga was 77.1%, 83.3% and 81.2%, respectively; counting all 450 non-responders as non-compliant gave conservative estimates of 53.4%, 57.7% and 56.3% for the three practices (n = 1,466). In the responding cohort, relief rose with the five-level compliance dose, from 5.2% (6/115) at dose 0 to 98.2% (326/332) at dose 4 (all three practices plus a repeat visit).

**Conclusions:** In this exploratory single-arm cohort, patient-reported relief was more frequent at higher levels of a composite measure that combined the three practices and repeat visits. Because there was no comparison group and all lifestyle components were delivered jointly alongside continuing pharmacotherapy, causality cannot be inferred. The findings support feasibility and justify prospective, controlled, disease-specific studies. The efficacy of the four-spice infusion remains under study and is not yet a proven treatment.

## Introduction

Non-communicable diseases (NCDs) threaten health gains, productivity and household budgets worldwide. India’s burden is large and rising: an estimated 101 million adults have diabetes, 315 million have hypertension and 254 million have generalised obesity [1].

Unhealthy diets, physical inactivity, tobacco use and excess body weight are the principal modifiable drivers [1–3]. In the Global Burden of Disease Study 2021, a substantial share of disability-adjusted life-years was attributable to these behavioural and metabolic risks, so a corresponding share of the burden is potentially avoidable through prevention [3]. Because these exposures are widespread and often coexist, prevention offers large health and economic returns.

The need is not confined to low- and middle-income countries. In the United States, chronic diseases are the leading causes of illness, disability and death and account for about 90% of the nation’s $5.3 trillion in annual health care expenditure; three in four adults have at least one chronic condition and more than half have two or more [4]. More than 40 million Americans have diabetes, a further 115 million adults have prediabetes, and obesity affects about 40% of adults [5]. Because the same modifiable behaviours — poor nutrition, physical inactivity, tobacco use and harmful alcohol use — drive this burden [4,5], lifestyle intervention is needed both to prevent disease and as an essential component of therapy alongside pharmacotherapy, in high-income as well as Indian settings [2,3].

Lifestyle interventions can prevent or improve NCDs. In Asian Indians with impaired glucose tolerance, lifestyle modification reduced incident diabetes by 28.5% relative to standard advice in the Indian Diabetes Prevention Programme [6]. Plant-based dietary patterns [7], Yoga for hypertension [8] and Yoga-based programmes for glycaemic control [9] have supportive evidence, and a 2026 meta-analysis of Indian adults with hypertension found that lifestyle interventions lowered systolic and diastolic blood pressure [10]. A multicentre Indian trial also found that a structured Yoga and lifestyle programme was feasible, with promising recruitment, follow-up and adherence in people at high risk of diabetes [11]. Much of this evidence, however, comes from efficacy trials or single-disease programmes. Less is known about whether a comprehensive, comprehensible lifestyle package that promotes self-determination can be delivered routinely alongside a busy family-medicine clinic, especially for multimorbid industrial workers [12,13]. Implementation is constrained by limited time, staffing, tools and financing among providers [14], and patients face barriers to sustained adherence [15].

ESIC Medical College & Hospital, Faridabad, India, provides free care at the point of use to industrial workers and their families covered by the Employees’ State Insurance Scheme (ESI Scheme). Within this tertiary-care setting, patients are, however, free to move between hospital departments. The Integrative Health Clinic (IHC) is a family-medicine-style outpatient clinic open to patients of all ages with chronic conditions. Care is organised around the patient’s context, beliefs and goals rather than a single diagnosis, and it explores and addresses the root causes of illness to support overall health and recovery. All patients receive the lifestyle intervention as an add-on to standard of care, aimed at integrative body–mind, whole-person care, with a focus on lifestyle change.

For this, it follows the 21-item Health Building Lifestyle Guideline, with a tracked 0–21 lifestyle score [2,16,17], and delivers care through a 7-step protocol based on the COM-B model [16]. The programme combines a plant-based diet, daily Yoga asanas and breath control (pranayama), and a home-prepared four-spice infusion [18–23]. All patients are followed up through calls, during which they are asked about their regularity with the three advised practices and any relief of presenting symptoms.

The clinic is led by a physician trained in public health and therapeutic Yoga, who demonstrates Yoga in the clinic with two on-site medical interns.

Prior single-disease signals from this clinic — an irritable bowel syndrome (IBS) remission case report combining diet, lifestyle and Yoga [19], a randomised trial of pranayama in exposed healthcare professionals [24] and public descriptions of the approach [25] — motivated a larger evaluation. We therefore examined a 20-month real-world cohort of 1,737 patients to assess feasibility, compliance with the three lifestyle components and patient-reported relief, and to determine whether the programme could be delivered within routine family-medicine care. We explicitly address the single-visit constraint through return-visits to the clinic and call-based feedback.

## Methods

### Design and aims

This was a retrospective single-arm cohort reported in accordance with the STROBE statement [26]. The study aimed to assess compliance with the three lifestyle components separately — spice infusion, diet and Yoga — and patient-reported relief. The analysis coded each outcome once per patient using the most recent definitive answer rule.

### Setting and dates

The observation window comprised the 20-month incident period from 1 February 2025 to 30 September 2026, with follow-up records through 4 November 2026. Data collection began in January 2025, but the January records were lost in a hospital system upgrade and are unavailable for this analysis. The clinic began in September 2024 as a pilot, initially functioning on Saturdays and was converted into a regular, six days a week (9 AM–1 PM) unit since 1 December 2024 under a Dean’s office directive. Hospital departments were advised to refer patients with chronic conditions, and patients also came by word of mouth. Our incident cohort was defined by the cut-off rule: a first visit on or before 30 September 2026. Records from October 2026 contributed follow-up data only, and patients whose first visit fell after 30 September 2026 (98 patients, 116 records) were excluded. The analysed denominator comprised patients with a usable telephone contact who were called; patients without a usable telephone contact were excluded from all outcome analyses; they are retained in the cohort flow and compared with the called group in Table 5.

### Medication posture

Prescribing at the IHC stayed within safe, guideline-concordant categories, with a conservative use of drugs; for example, topical non-steroidal anti-inflammatory drugs (NSAIDs) were preferred to oral tablets for musculoskeletal pain; antitussives were discouraged while steam inhalation was advised for cough; antacids were discouraged in favour of a plant-based diet for dyspepsia; specific drugs for constipation were avoided in favour of a whole-food, plant-based diet, adequate hydration and movement. Given the widespread prevalence of vitamin-mineral deficiency in India, and particularly among the lower socioeconomic groups, to which most the clinic’s target population belonged, vitamin and mineral supplements were liberally prescribed. No medication-related adverse effects were recorded during the observation period.

### Participants

and diagnosis coding. Diagnoses were grouped into 14 systems and 323 canonical conditions for burden counting after mapping 699 raw diagnosis labels. Although patients could have conditions in multiple systems, each patient was analysed as a single unit, with outcomes counted once per patient, not per episode or visit. Insurance numbers, where present, were used solely for the family-linkage analysis and do not appear in any patient-level output.

### Intervention (replicable)

Within this care model, advice on three lifestyle components was added to standard pharmacotherapy. The first was a home-prepared morning infusion of four whole culinary spices: 10 g each of coriander (Coriandrum sativum) seed, fenugreek (Trigonella foenum-graecum) seed, cumin (Cuminum cyminum) seed and cinnamon (Cinnamomum spp.) bark, soaked overnight in 1 litre of water. A glassful was taken lukewarm first thing in the morning before Yoga; the water was replenished for up to three days, after which a fresh infusion was prepared. The infusion was advised as one component of the clinic’s three-component lifestyle programme, alongside dietary modification and daily Yoga, and was never evaluated separately. Patients were advised to continue prescribed pharmacotherapy. The second was a plant-based diet and diurnal routine based on the 21-item Health Building Lifestyle Guideline [16], including red/green/yellow items, motivational interviewing and team reinforcement. The third was daily practice of 20 Yoga asanas and pranayama: the physician demonstrates the sequence in the clinic, patients practise under supervision, and a video-series link is shared to support home practice [18]. The guideline was synthesised from WHO and ICMR-NIN guidance and behaviour-change principles (COM-B, Health Belief Model and self-efficacy) [2,16,17] and delivered through its published 7-step protocol (detailed sequence in Appendix A): lifestyle assessment; traffic-light education; self-scored baseline; barriers and goal-setting; physician reinforcement; physician-led demonstration; and follow-up calls [16]. Group medical attendance, and whole family care, where feasible, were encouraged.

### Clinic workflow and time per patient

The clinic was organised around a zero-waiting workflow in a single room with Yoga mats. Before their consultation, patients read the bilingual lifestyle guideline [27] and glanced through Yoga and lifestyle web content shared on WhatsApp on their own mobile. They also observed repeat-visit patients by their side practising Yoga and pranayama and, intermittently, heard them recounting their experiences. A trainee doctor or nurse then saw patients one at a time for vital signs, anthropometry and history as a preliminary assessment. Thereafter, the clinic doctor examined each patient to establish the diagnosis. Standard care, including clinically indicated investigations, was provided for these diagnoses. Patients returned to the mat to practise Yoga and pranayama while prescriptions and the investigation list were prepared. Before leaving, lifestyle advice was repeated and recall done; patients were asked to follow-up in 2–3 weeks with test reports. At the repeat visits, compliance was reviewed and patients’ experiences heard; weight was rechecked as a marker of compliance; goals were revisited and advice was reinforced in person, by telephone or by WhatsApp. Each patient spent about 30 minutes per visit over sessions that covered knowledge, attitude and supervised practice of the asanas. Such an elaborate protocol aimed to operationalised the three COM elements — Capability, Opportunity and Motivation — so that patients leave the clinic with a concrete inventory of lifestyle changes to be followed and, with the clinic’s experience, return with a sense of motivation and empowerment to take responsibility for their own health.

### Outcomes

The primary outcome was the patient-level coded classification of reported relief (YES/NO/no definitive answer). Secondary outcomes were the patient-level compliance of spice, diet and Yoga practices. Records were ordered by call date, and the most recent definitive answer (YES/NO) was taken as the patient’s code; if no definitive answer was recorded (“no response” or “not interested”), the patient was classified as having no definitive answer. “Not interested” was treated as an intentional non-answer, not a definitive NO.

### Relief measurement

Relief was deliberately elicited with a single generic question. The term “Relief” is open to individual interpretations and may reflect the patient’s most pressing problems rather than a standardised, disease-specific construct. Validated disease-specific questionnaires would have been preferable, but they were impractical in a large, diagnostically diverse cohort. A single item of :”relief” was chosen for simplicity, comprehensibility, flexibility across conditions and acceptability to patients. For future rounds, we propose adding a percentage-relief question — “compared with your first visit, what percentage relief have you had?” — where 0 represents no relief, to improve discrimination.

### Dose levels

The five-level compliance with the three lifestyle practices was defined as follows: 0 = no components, 1 = any one component and 2 = any two components of the three (spice, diet and Yoga); 3 = all three components compliant; and 4 = all three components compliant with at least one revisit. A revisit alone never qualified as a dose. Dose–response is reported as two estimates: a responder estimate (YES among patients with a definitive answer, YES/NO) and a conservative estimate (YES among all analysed patients, including patients with no definitive answer in the denominator).

### Response attribution

All patients with a usable telephone contact were contacted. “Contacted” means that a call date was recorded at any point (800 of 1,035 single-visit and 666 of 702 revisit patients). Because diet, Yoga, the infusion and medicines were delivered jointly, attribution to any single component was not possible.

### Screening definitions

Screening operated at two levels. Clinical screening meant identification of behavioural risks and other conditions from history and examination — for example, tobacco abuse — and their addition to the coded diagnosis; all the 114 tobacco-abuse diagnoses (114 patients; tobacco abuse is a single canonical condition, so pairs = patients) were identified in this way. Objective-evidence screening was applied to valid records for conditions that could be unobserved or silent: obesity-suggestive (BMI >28 kg/m² without an obesity code on the same record), diabetes-suggestive (HbA1c ≥6.5% without a diabetes code), sustained unlabelled hypertension (≥2 readings with systolic ≥140 mmHg or diastolic ≥90 mmHg on records without a hypertension label) and anaemia-suggestive (haemoglobin <10 g/dL without an anaemia code). A flagged patient was added to the main counts only if they had never been coded with that condition anywhere in the record; patients whose code appeared at another visit were excluded. Thresholds were pragmatic single-cut choices. Because the hypertension rule requires two elevated readings on separate visits, all 39 never-coded hypertension patients had at least one revisit; the obesity, diabetes and anaemia screens, which require only a single measurement, also identified single-visit patients. Alternative cut-off checks increased the never-coded counts without changing the overall pattern: obesity 161 → 198 at BMI ≥27.5 and 415 at BMI ≥25; anaemia 44 → 264 with WHO adult haemoglobin thresholds (<13 g/dL in men, <12 g/dL in women). The 246 objective-evidence additions are reported in the Table 2 footnote and Table S3; clinically screened diagnoses such as tobacco abuse are included in the main counts but are not part of the 246 objective-evidence additions.

### Ascertainment policy

The main burden tables include all patient–condition pairs from contacted patients. Clinically screened diagnoses were added to the coded record. Table 2 and Table S3 report the 246 objective additions and their provenance; this avoids under-counting objective disease while retaining the coded diagnosis as the clinical record.

### Harms

No harms were reported in this retrospective extraction. Concurrent medicines were continued throughout. As this was a record review without prospective safety monitoring, the absence of recorded harms refers to documented events only.

### Statistics

Analyses were descriptive, with explicit denominators and column percentages. Classification was once per patient by the most recent definitive answer, and correlations used analysed-only denominators. The primary relief estimate used the 1,016 analysed patients with a definitive answer. The conservative lower bound used all 1,466 analysed patients as the denominator and counted every non-YES (NO and no definitive answer) as no relief. We did not assume relief (or lack of it) for non-responders and therefore did not calculate a best (or worst) case estimate. Disease-specific paired analyses with effect sizes, confidence intervals and sensitivity analyses are deferred to follow-up papers, one disease at a time (for example, hypertension, diabetes, obesity, arthritis or acid-peptic disease).

### Ethics and approval

The retrospective, de-identified case-series analysis was notified to the Institutional Ethics Committee (IEC), ESIC Medical College & Hospital, Faridabad (EC Reg. No. EC/NEW/INST/2023/HR/0307), on 2 March 2026. The Committee in its letter of 18 March 2026 raised no objection to publication, provided that confidentiality and anonymity were strictly maintained and no identifiable information was disclosed. On 22 October 2026, the Committee extended its no-objection to the proposed analysis and to matching family members by insurance number for analytical purposes, subject to the same conditions of strict confidentiality and anonymity, non-disclosure of identifiable information and compliance with applicable institutional policies. The no-objection determinations cover the de-identified data reported here, including follow-up records through 4 November 2026. Patients younger than 18 years were dependants of insured workers and were included as part of the same de-identified record review.

## Results

### Flow

The incident-window rule excluded 98 patients (116 records) whose first visit fell after 30 September 2026, and the validity filter then excluded 183 records without a coded diagnosis entry (fully undiagnosed patients and stray blank rows), leaving 4,047 valid records from 1,737 unique patients (1,035 single-visit and 702 revisit patients). Sex and age were available for all 1,737 patients; outcome data only for the 1,466 called. Per-dose denominators are given in Table 4, and monthly valid-diagnosis records ranged from 91 to 269 (all cleaned records in Figure 1 ranged from 93 to 280). Cohort flow and baseline characteristics are summarised in Table 1, and the two contact groups are compared in Table 5.

**Figure 1.**
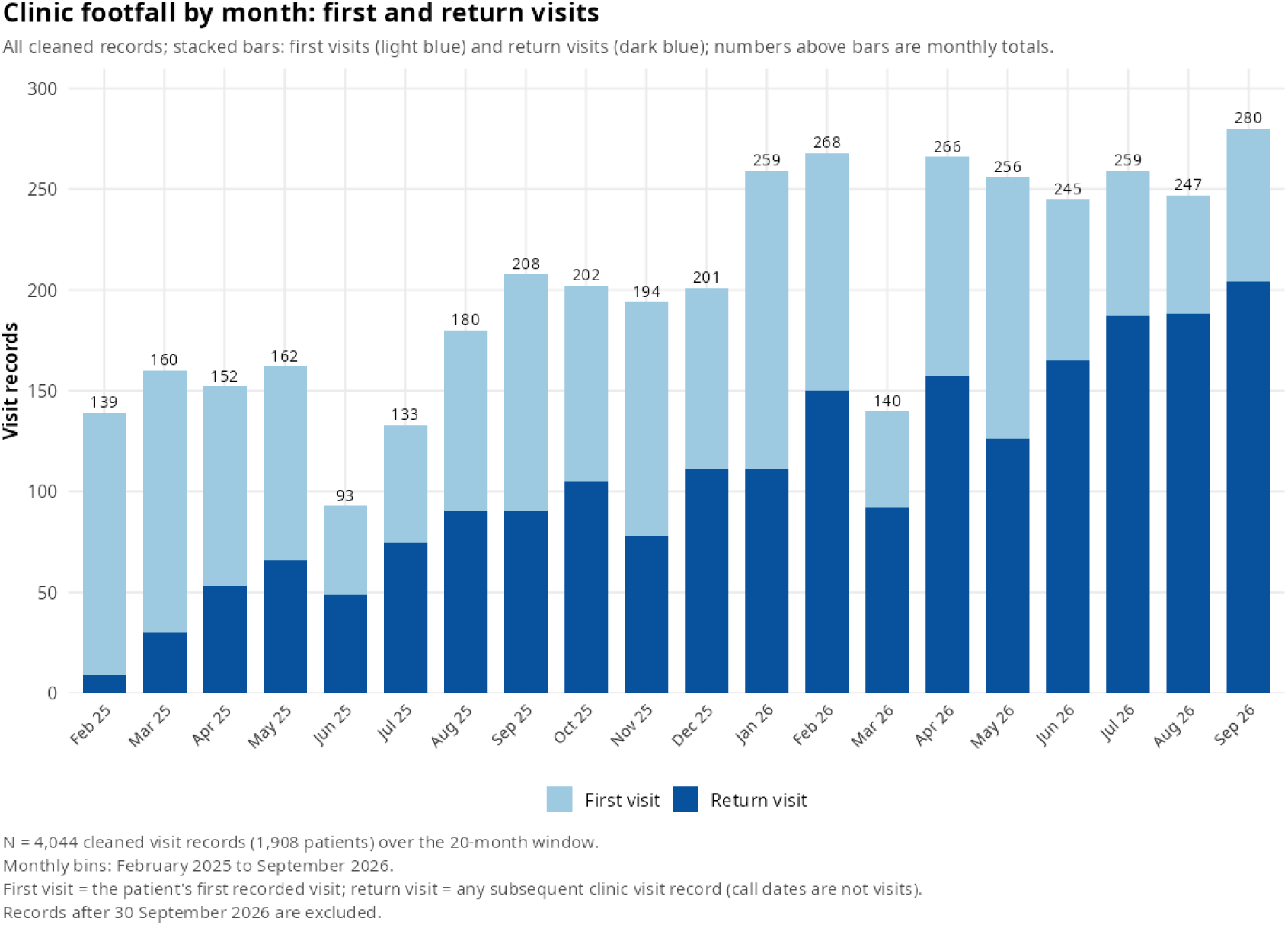
Clinic footfall by month: first and return visits. All cleaned records in the 20-month window; stacked bars: first visits (light blue) and return visits (dark blue); numbers above bars are monthly totals. N = 4,044 cleaned visit records in the 20-month window (1,908 patients); records after 30 September 2026 are excluded. Of the 1,908 patients, 1,737 had at least one coded diagnosis and form the analysis cohort; the remaining 171 had no coded diagnosis and are excluded from all burden and outcome tables. Monthly totals ranged from 93 (June 2025) to 280 (September 2026); monthly valid-diagnosis records ranged from 91 to 269. Monthly bins: February 2025 to September 2026 (20 months). First visit = the patient’s first recorded visit; return visit = any subsequent clinic visit record (call dates are not visits).

**Table 1.** Cohort flow and baseline characteristics.

| Characteristic | Value |
|---|---|
| Cleaned visit records (valid UHID and visit date) | 4,346 |
| Records excluded: first visit after 30 September 2026 (98 patients; 116 records) | 116 |
| Records excluded: no coded diagnosis (blank or summary rows) | 183 |
| Valid records | 4,047 |
| Unique patients | 1,737 |
| Female | 904 (52.0%) |
| Female age, mean ± SD (years) | 40.3 ± 14.6 |
| Male | 833 (48.0%) |
| Male age, mean ± SD (years) | 44.8 ± 15.1 |
| Single-visit patients | 1,035 (59.6%) |
| Revisit patients | 702 (40.4%) |
| Visits per patient, mean | 2.33 |
| Usable telephone contact and called (analysis denominator) | 1,466 (84.4%) |

Values are counts of records or patients; percentages are of the 1,737-patient cohort unless otherwise stated. A record was valid when at least one diagnosis field contained a coded entry; empty, “NA” and “0” entries were excluded. The incident cohort was defined by a first visit on or before 30 September 2026; records after that date are follow-up only. Disease-wise analyses were restricted to the 1,466 patients with a usable telephone contact who were called; the 271 patients without one are compared with them in Table 5. Figure 1 plots the 4,044 cleaned records dated on or before 30 September 2026 (1,908 patients); the 4,047 valid records here additionally include 184 follow-up visits recorded after that date for patients whose first visit preceded the cut-off, and exclude 183 records without a coded diagnosis (181 of them within the Figure 1 window).

### Baseline characteristics and burden

The cohort of 1,737 comprised 904 female patients (52.0%; mean age 40.3 ± 14.6 years) and 833 male patients (48.0%; mean age 44.8 ± 15.1 years). The analysis was restricted to the 1,466 patients with a usable telephone contact who were called. System-level sex, age and revisit distributions for the analysed patients are given in Table S1. Among them, the most frequent patient–condition pairs were hypertension (511), obesity (397), arthritis (268), diabetes mellitus (246), gastritis (163), tobacco abuse (114), hypothyroidism (103) and anaemia (99); complete system-level case-loads are shown in Table 2. Tobacco abuse was the most frequent behavioural-risk code: 114 patients (114 patient–condition pairs; 7.8% of the analysed cohort; 16.2% of the 703 men), all male. All tobacco-abuse cases were identified through clinical screening (history and examination) and added to the diagnosis. Objective-evidence screening contributed 246 patient–condition pairs in 233 patients, with 13 patients meeting criteria for two conditions (obesity 161, hypertension 39, anaemia 44, diabetes 2); these pairs are included in the Table 2 counts and detailed in the Table 2 footnote and Table S3. In the full 1,737-patient cohort, coded-diagnosis prevalence was hypertension 29.8% (518), arthritis 18.0% (313), diabetes mellitus 15.6% (271), obesity 15.0% (260) and gastritis 11.1% (193); these are coded diagnoses in the full cohort and differ from the patient–condition pair counts in Table 2, which include screening-detected cases among the 1,466 called patients. Patients without a usable telephone contact (n = 271) were similar in sex and age but were more often single-visit (86.7% vs 54.6%) and had a lighter cardiometabolic case-mix (hypertension 17.0% vs 32.2%, obesity 8.9% vs 16.1%, diabetes 10.0% vs 16.6%; Table 5). Because these patients had fewer and milder recorded conditions, their exclusion would, if anything, tend to understate rather than inflate the benefit observed; this remains an inference, as their outcomes were not measured.

**Table 2.** Disease-system case-load (patient–condition pairs)

| System | Patients | Patient–condition pairs | Mean distinct conditions per patient | Most frequent conditions (n) |
|---|---|---|---|---|
| ENDOCRINE / METABOLIC | 636 | 815 | 1.28 | Obesity 397; Diabetes Mellitus 246; Hypothyroidism 103; Fatty Liver 25; Dyslipidemia 9; Growth Retardation 5; Bronzed Diabetes 4; Hyperthyroidism 4 |
| CARDIOVASCULAR | 535 | 548 | 1.02 | Hypertension 511; Varicose Veins 10; Coronary Artery Disease 6; Angina Pectoris 4; Cardiac Pacemaker Syndrome 2; Buergers Disease 1; Cardiac Asthma 1; Cardiac Disorder 1 |
| MUSCULOSKELETAL | 479 | 539 | 1.13 | Arthritis 268; Spondylitis 90; Sciatica 53; Cervical Spondylitis 17; Osteoarthritis 14; Musculoskeletal Disorder 13; Gout 10; Rheumatoid Arthritis 8 |
| GASTROINTESTINAL | 340 | 381 | 1.12 | Gastritis 163; Hemorrhoids 68; Constipation 43; Gastric Ulcer 20; Irritable Bowel Syndrome 9; Oesophagitis 8; Anal Fissure 6; Cholelithiasis 6 |
| BEHAVIORAL / MENTAL | 167 | 172 | 1.03 | Tobacco Abuse 114; Insomnia 23; Anxiety Disorder 11; Anorexia Nervosa 5; Depression 5; Alcohol Abuse 3; Alcoholism 2; Alcohol Dependence 1 |
| GYNAE / REPRODUCTIVE | 162 | 188 | 1.16 | Dysmenorrhea 45; Pelvic Inflammatory Disease 29; Polycystic Ovary Syndrome 22; Uterine Fibroid 12; Ovarian Cyst 10; Female Infertility 9; Male Infertility 6; Menstrual Disorder 5 |
| DIETARY / NUTRITIONAL | 161 | 171 | 1.06 | Anemia 99; Vitamin Deficiency 63; Malnutrition 3; Vitamin D Deficiency 3; Nutritional Disorder 1; Nutritional Wasting 1; Vitamin C Deficiency 1 |
| RESPIRATORY | 118 | 132 | 1.12 | Asthma 42; Rhinitis 38; Bronchitis 17; Chronic Obstructive Pulmonary Disease 15; Upper Respiratory Tract Infection 7; Allergic Rhinitis 4; Chronic Rhinitis 3; Allergic Bronchopulmonary Aspergillosis 1 |
| NEUROLOGICAL | 89 | 101 | 1.13 | Migraine 47; Daily Headache 8; Headache 7; Cluster Headache 5; Epilepsy 4; Cerebral Decompression Injury 2; Hemiparesis 2; Hemiplegia 2 |
| RENAL / UROLOGY | 81 | 89 | 1.10 | Urinary Tract Infection 31; Renal Stone 16; Benign Prostatic Hyperplasia 9; Chronic Kidney Disease 8; Kidney Disease 3; Urinary Incontinence 3; Hydrocele 2; Hydronephrosis 2 |
| INFECTIONS | 61 | 62 | 1.02 | Worm Infestation 16; Amoebiasis 10; Taeniasis 6; Tuberculosis 6; Typhoid Fever 5; Malaria 3; Gonorrhea 2; Sexually Transmitted Disease 2 |
| SKIN | 47 | 53 | 1.13 | Eczema 10; Urticaria 5; Allergic Dermatitis 4; Vitiligo 4; Dermatitis 3; Psoriasis 3; Skin Allergy 3; Acne Vulgaris 2 |
| NEOPLASTIC / CYSTIC | 15 | 16 | 1.07 | Fibromatosis 3; Neurofibromatosis 2; Tumor Of Urinary Tract 2; Adenocarcinoma 1; Breast Tumor 1; Cancer Metastatic To Gallbladder 1; Cervical Cancer 1; Familial Multiple Lipomatosis 1 |
| EYE / ENT | 12 | 13 | 1.08 | Sinusitis 3; Adenoid Hypertrophy 1; Epistaxis 1; Eye Infection 1; Hyperopia 1; Impacted Cerumen 1; Lymphadenopathy 1; Otitis Externa 1 |

Counts are for the 1,466 analysed patients. Conditions are free-text diagnoses mapped to 14 systems (699 distinct raw labels; 323 canonical conditions in this cohort); all labels were mapped to a system. “Patients” counts distinct patients with at least one condition in the system, and patients with conditions in multiple systems appear in each such system’s row. “Patient–condition pairs” counts distinct patient–condition combinations (each patient counted once per condition), and the mean is the number of pairs divided by the number of patients; the n in “Most frequent conditions” is patient–condition pairs. Clinical screening (history and examination) identified behavioural risks such as tobacco abuse; these diagnoses are included in the main counts but are not part of the totals reported for the screening contribution. Objective-evidence screening contributed 246 pairs in 233 patients (obesity 161, hypertension 39, anaemia 44, diabetes 2; 13 patients had two screening-detected conditions). Thresholds were BMI >28 kg/m<sup>2</sup>, HbA1c ≥6.5%, ≥2 blood-pressure readings with systolic ≥140 mmHg or diastolic ≥90 mmHg, and haemoglobin <10 g/dL; condition-level provenance is in Table S3. Column totals: 3,280 pairs across 14 systems.

### Clinic uptake and continuity

Clinic footfall grew over the observation window, from 139 recorded visits in February 2025 to 280 in September 2026. The net growth came from returning patients: first visits fell from 130 in February 2025 to 76 in September 2026 (59 in August), while return visits rose from 9 to 204 (188 in August). The return-visit share of monthly visits therefore climbed from 6.5% to 72.9% (72–76% in July–September 2026; Figure 1); part of this rise is mechanical, because the cohort started empty and accumulated revisits over time. Participation also spread within families. Grouping patients by shared insurance number (the insured worker and registered dependants share one number) yielded 1,412 families, of which 274 (19.4%) were represented by more than one member. In 138 of these families (50.4%), at least one member first attended after the family’s first member (180 patients, 10.4% of the cohort; median interval 64.0 days, interquartile range (IQR) 18.5–195.5); the remaining 136 multi-member families attended on the same day. Patients spanned all ages, from infancy to 87 years; 101 patients (5.8%) were under 18, including 40 children among the 180 who attended after a family member. Together with the return-visit pattern, this family clustering is consistent with gradual intra-family diffusion of the programme (a clinic observation; referral pathways were not audited).

### Provider referrals

Conventional-medicine providers regularly referred chronic, resistent cases to the clinic, notably from Obstetrics & Gynaecology, Internal Medicine, Gastroenterology and Psychiatry. The nature of referral patterns were a clinic observation and were not systematically audited; they may contribute to a case mix weighted toward chronic, previously managed conditions.

### Contact, outcomes and adherence

Of the 1,466 analysed patients, 1,016 answered the practice items and gave a definitive relief answer. Of the 1,466 analysed patients, 850 (83.7%; 95% CI 81.3–85.8) reported relief (YES) and 166 reported no relief (NO); the remaining 450 (850 + 166 + 450 = 1,466) gave no definitive answer. Treating all 450 non-responders as no relief, the conservative lower bound of reported relief was 58.0% (850/1,466). The 26 patients recorded as “not interested” were retained as having no definitive answer, not NO, and were not counted as relief. All 1,016 patients with a definitive relief answer also answered the three practice items on the same call. Compliance was YES for the spice infusion in 783 patients (77.1%), for diet in 846 (83.3%) and for Yoga in 825 (81.2%) (Table 3). Counting all non-responders as non-compliant gave lower bounds of 53.4%, 57.7% and 56.3% (n = 1,466). Among the 86 patients with tobacco abuse and a codable relief vote, 77 (89.5%) reported relief. Non-responders were not assumed to have relief (or lack of it), so no best (or worst) case estimate is reported.

**Table 3.**
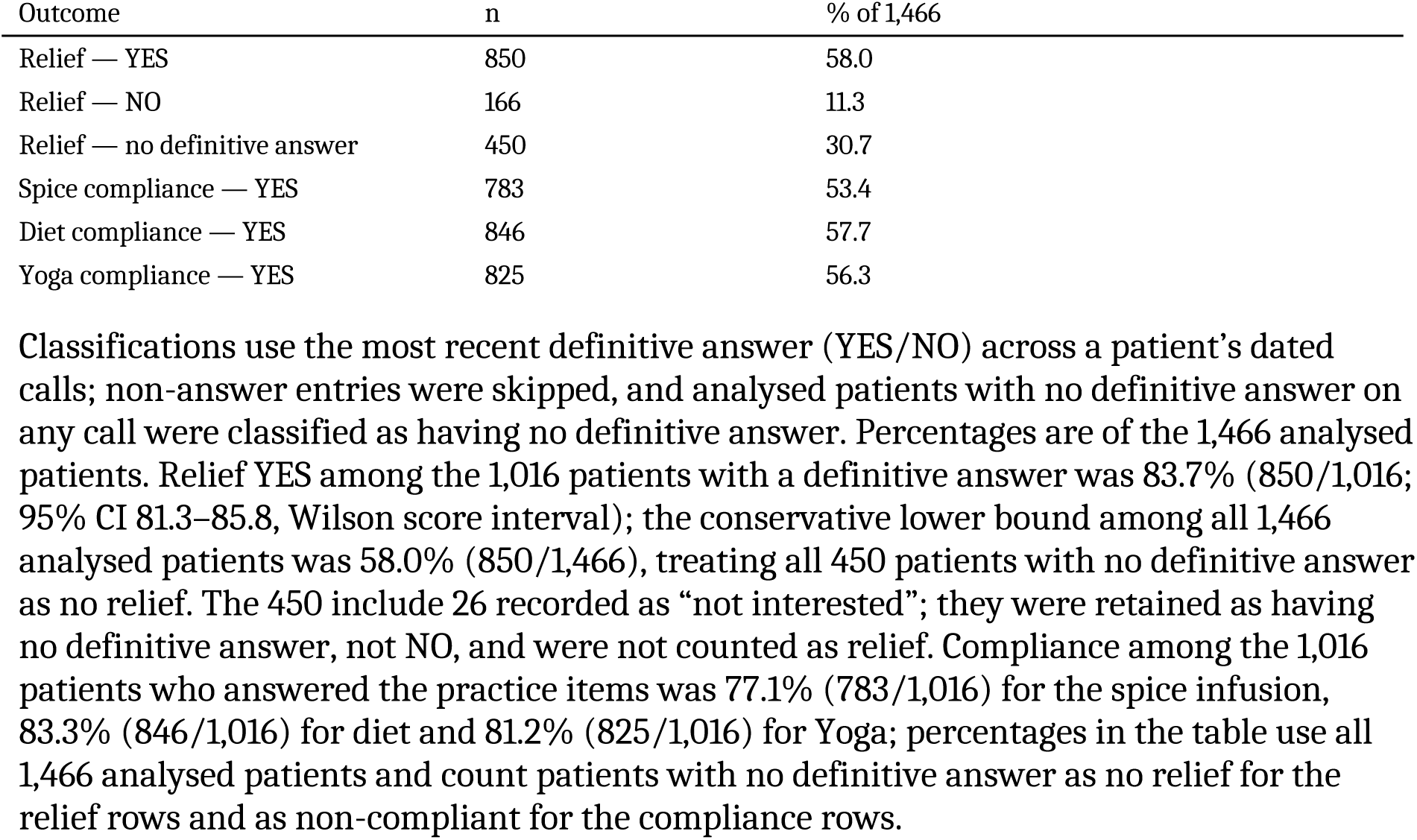
Patient-level outcomes among analysed patients (n = 1,466)

| Outcome | n | % of 1,466 |
|---|---|---|
| Relief — YES | 850 | 58.0 |
| Relief — NO | 166 | 11.3 |
| Relief — no definitive answer | 450 | 30.7 |
| Spice compliance — YES | 783 | 53.4 |
| Diet compliance — YES | 846 | 57.7 |
| Yoga compliance — YES | 825 | 56.3 |

Classifications use the most recent definitive answer (YES/NO) across a patient’s dated calls; non-answer entries were skipped, and analysed patients with no definitive answer on any call were classified as having no definitive answer. Percentages are of the 1,466 analysed patients. Relief YES among the 1,016 patients with a definitive answer was 83.7% (850/1,016; 95% CI 81.3–85.8, Wilson score interval); the conservative lower bound among all 1,466 analysed patients was 58.0% (850/1,466), treating all 450 patients with no definitive answer as no relief. The 450 include 26 recorded as “not interested”; they were retained as having no definitive answer, not NO, and were not counted as relief. Compliance among the 1,016 patients who answered the practice items was 77.1% (783/1,016) for the spice infusion, 83.3% (846/1,016) for diet and 81.2% (825/1,016) for Yoga; percentages in the table use all 1,466 analysed patients and count patients with no definitive answer as no relief for the relief rows and as non-compliant for the compliance rows.

### Full-trio practitioners

A total of 697 patients (47.5% of the 1,466 analysed) practised all three components (spice infusion, diet and Yoga): of these, 365 had a single visit and 332 had at least one revisit. Reported relief was YES in 679 (97.4%; Table 4, Figure 2). Full-trio patients accounted for 679 of the 850 relief reports in the analysed cohort (79.9%). Relief was reported by 165 of 204 patients practising one or two components (80.9%) and by 6 of 115 respondents practising none (5.2%; respondents only). Relief was found to be dose-gradient dependent (Table 4): 326/332 (98.2%) at dose 4, 353/365 (96.7%) at dose 3, 142/159 (89.3%) at dose 2, 23/45 (51.1%) at dose 1 and 6/115 (5.2%) at dose 0 among respondents, with the conservative estimate identical at doses 1–4 and 1.1% (6/565) at dose 0 (Figure 2). Among analysed patients (n = 1,466), the correlation between full-trio compliance and relief was 0.55 (phi); full-trio compliance was recorded in 45.6% of single-visit and 49.8% of revisit patients.

**Table 4.** Relief by compliance dose (responder and conservative estimates)

| Dose | Definition | Analysed | Answered | Relief YES | Responder estimate, % (YES/answered) | Conservative estimate, % (YES/analysed) |
|---|---|---|---|---|---|---|
| 0 | No component compliant (includes 450 with no compliance data; see note) | 565 | 115 | 6 | 5.2 | 1.1 |
| 1 | Any one component compliant | 45 | 45 | 23 | 51.1 | 51.1 |
| 2 | Any two components compliant | 159 | 159 | 142 | 89.3 | 89.3 |
| 3 | All three components compliant, single visit | 365 | 365 | 353 | 96.7 | 96.7 |
| 4 | All three components compliant plus revisit | 332 | 332 | 326 | 98.2 | 98.2 |

Components are the spice infusion, diet and Yoga, each coded per patient under the most recent definitive answer rule. Dose 4 requires all three components plus at least one revisit; a revisit alone never scores as a dose. The responder estimate is the proportion with relief YES among patients who answered at that dose (patients with no definitive answer excluded); the conservative estimate uses all analysed patients at that dose as the denominator and counts every patient without a definitive answer as no relief. At doses 1–4 all analysed patients answered, so the two estimates coincide. At dose 0, 450 of the 565 analysed patients had no compliance data; the 1.1% (6/565) is a conservative floor over all analysed patients at this dose, not an estimate of relief among confirmed non-practitioners, for whom the responder estimate is 5.2% (6/115). Full-trio practitioners (n = 697) reported relief in 679 (97.4%), accounting for 79.9% of all relief reports. See Figure 2. Dose 0 is a default category, not confirmed non-adoption: 450 of the 565 patients at this dose had no compliance data and were assigned dose 0 because a practice without a YES is counted as not adopted; these patients also have no definitive relief answer.

**Table 5.** Comparison of patients called for follow-up and patients without a usable telephone contact.

| **Characteristic** | **Called (n = 1,466)** | **Not called (n = 271)** | **All (n = 1,737)** |
|---|---|---|---|
| Male | 703 (47.9%) | 130 (48.0%) | 833 (48.0%) |
| Female | 763 (52.1%) | 141 (52.0%) | 904 (52.0%) |
| Age, mean ± SD (years) | 42.4 ± 15.2 | 43.0 ± 14.0 | 42.5 ± 15.0 |
| Age <18 years | 89 (6.1%) | 12 (4.4%) | 101 (5.8%) |
| Single-visit | 800 (54.6%) | 235 (86.7%) | 1,035 (59.6%) |
| Revisit | 666 (45.4%) | 36 (13.3%) | 702 (40.4%) |
| **Most common conditions, n (% of group)** |  |  |  |
| Hypertension | 472 (32.2%) | 46 (17.0%) | 518 (29.8%) |
| Arthritis | 268 (18.3%) | 45 (16.6%) | 313 (18.0%) |
| Diabetes mellitus | 244 (16.6%) | 27 (10.0%) | 271 (15.6%) |
| Obesity | 236 (16.1%) | 24 (8.9%) | 260 (15.0%) |
| Gastritis | 163 (11.1%) | 30 (11.1%) | 193 (11.1%) |
| Tobacco abuse | 114 (7.8%) | 6 (2.2%) | 120 (6.9%) |
| Hypothyroidism | 103 (7.0%) | 17 (6.3%) | 120 (6.9%) |
| Spondylitis | 90 (6.1%) | 10 (3.7%) | 100 (5.8%) |
| Haemorrhoids | 68 (4.6%) | 8 (3.0%) | 76 (4.4%) |
| Vitamin deficiency | 63 (4.3%) | 9 (3.3%) | 72 (4.1%) |

Coded diagnoses only, using the same canonical diagnosis map as Table 2; the 246 objective-evidence screening additions reported in Table 2 are not counted here so that differences reflect recorded case-mix rather than measurement intensity (patients without a usable telephone contact were more often single-visit and had fewer blood-pressure, haemoglobin and BMI measurements). The patients themselves remain in the cohort and in the denominators. Denominators are patients in each group, not patient–condition pairs. Conditions are listed in order of overall frequency. The two groups were similar in sex and age, but patients not called were more often single-visit and had a lighter cardiometabolic case-mix.

**Figure 2.**
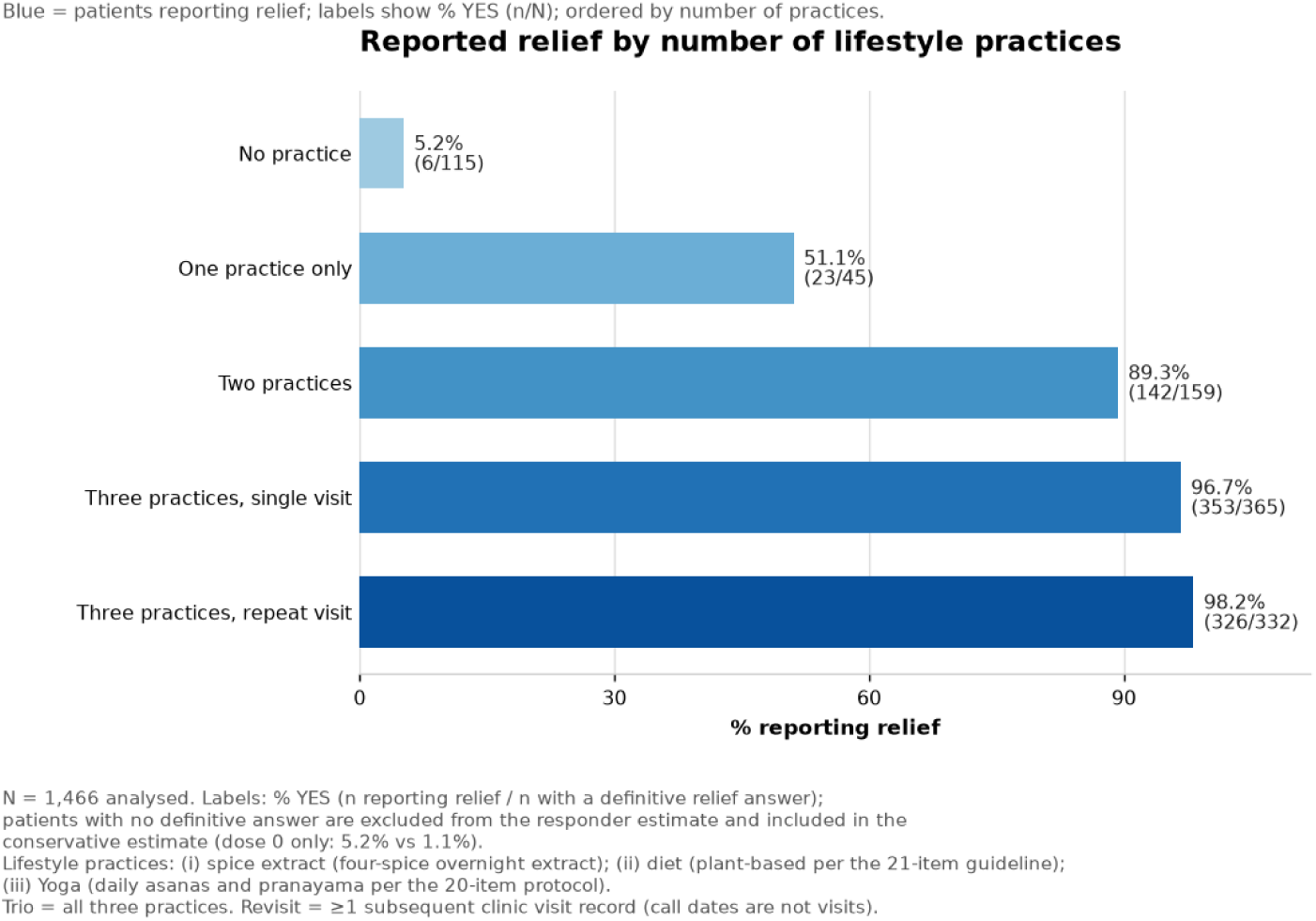
Reported relief by number of lifestyle practices. Bars, top to bottom: no practice; one practice only; two practices; all three practices with a single visit; all three practices with a repeat visit. Labels give the responder estimate as YES % (n reporting relief / n with a definitive relief answer at that dose); patients with no definitive answer are excluded from this estimate and included in the conservative estimate (dose 0 only: 5.2% vs 1.1%). N = 1,466 analysed; source counts in Table 4.

### Disease-system adherence and relief

Adherence and relief varied modestly across the 14 disease systems (Figures 3 and 4). Among analysed patients who answered, spice-infusion compliance ranged from 62.5% (eye/ENT) to 88.9% (skin), diet advice from 72.4% (neurological) to 95.0% (infections), and Yoga practice from 75.0% (eye/ENT) to 85.7% (neoplastic/cystic). Relief ranged from 75.0% (eye/ENT) to 89.7% (infections, 35/39). Several extremes arose in the smallest systems, and patients with conditions in more than one system are counted in each (Table 2), so these comparisons remain descriptive.

**Figure 3.**
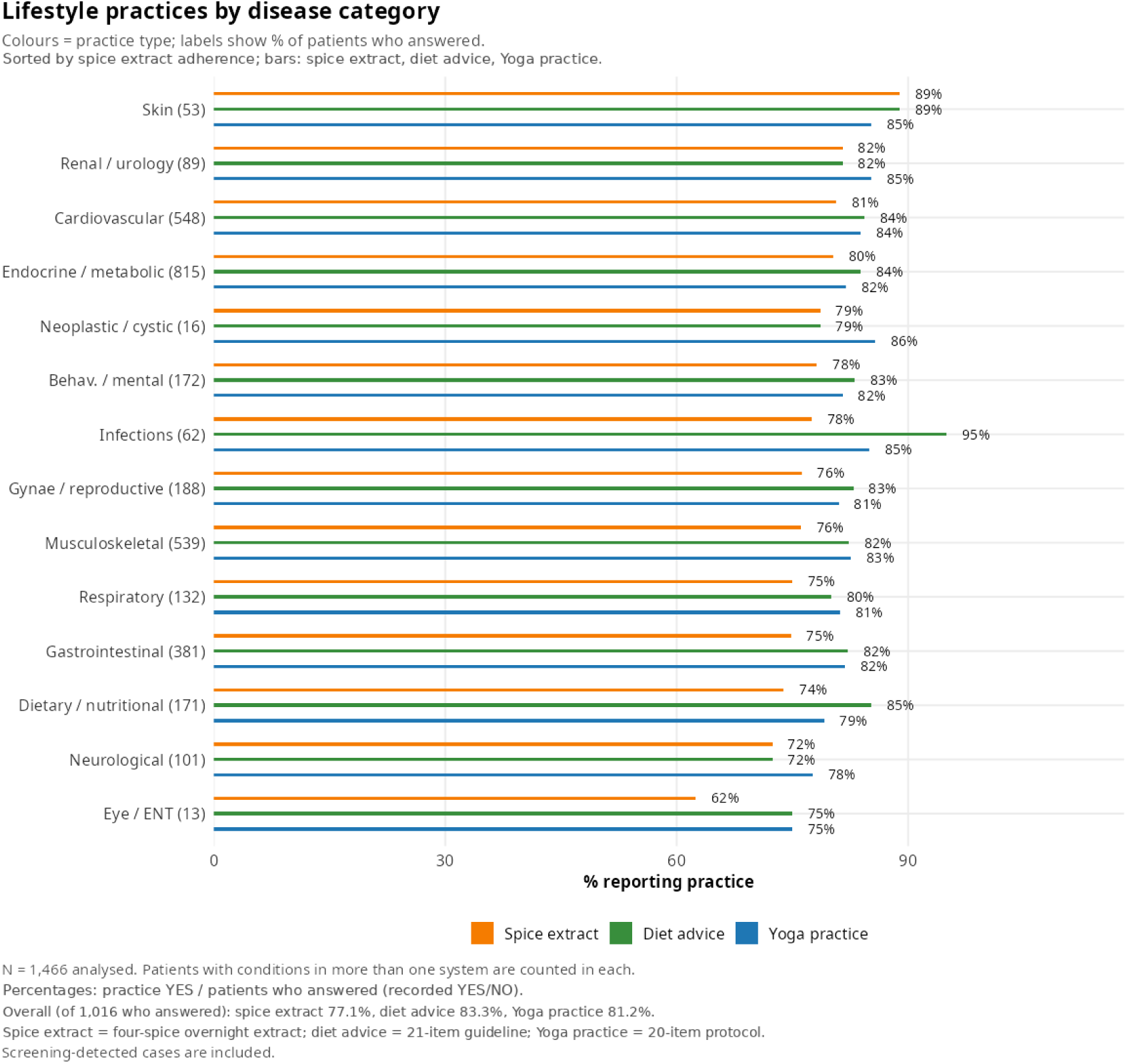
Lifestyle practices by disease category. Bars per system (patient–condition pairs in brackets), coloured by practice: spice infusion, diet advice, Yoga practice; systems sorted by spice-infusion adherence. Percentages are practice YES among analysed patients who answered (recorded YES/NO). Overall, among the 1,016 patients who answered: spice infusion 77.1%, diet advice 83.3%, Yoga practice 81.2%. Screening-detected cases are included; patients with conditions in more than one system are counted in each. N = 1,466 analysed.

**Figure 4.**
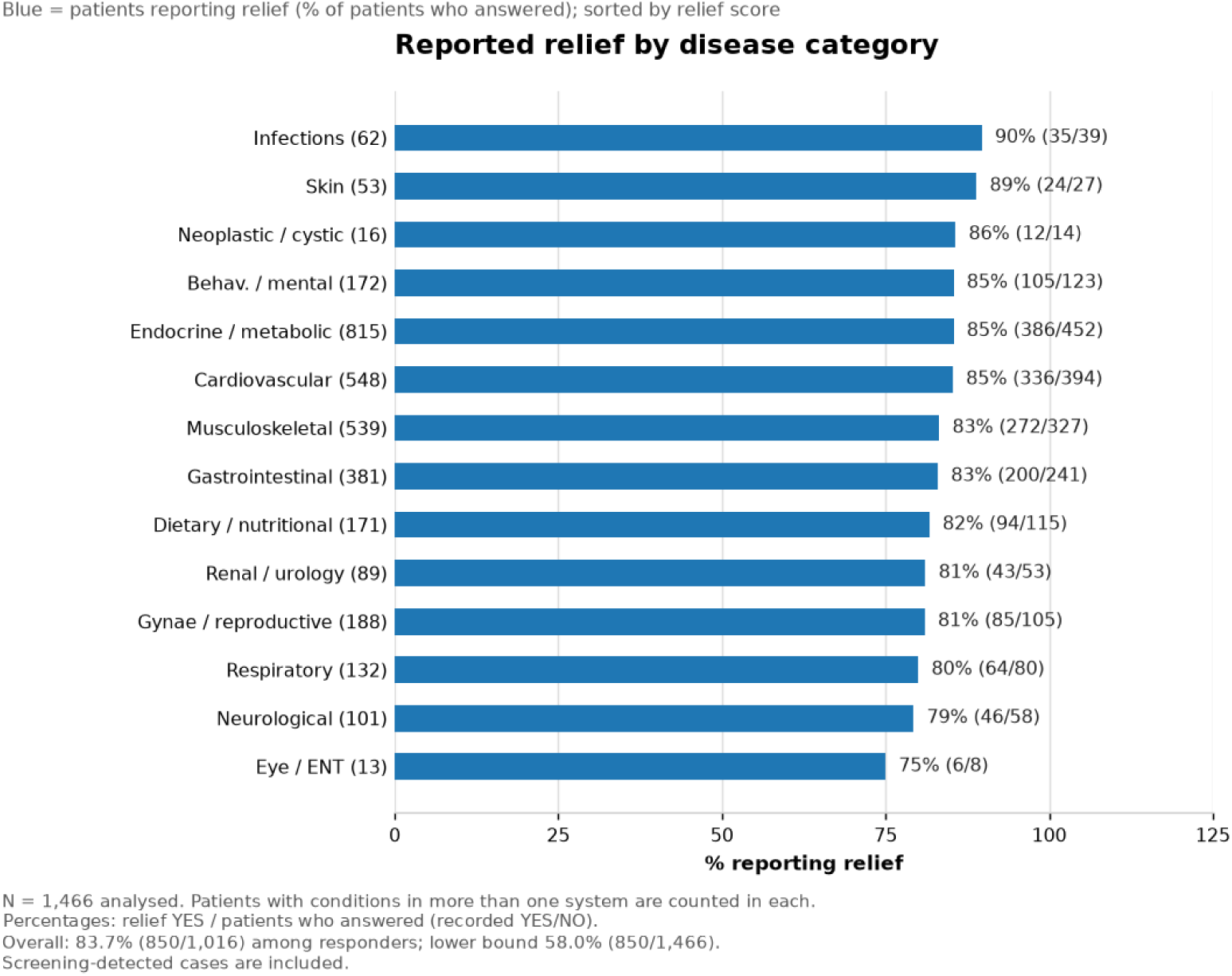
Reported relief by disease category. Bars: relief YES among analysed patients who answered; labels give the % with n/N; patient–condition pairs in brackets; systems sorted by relief. Overall: 83.7% (850/1,016) among responders; conservative lower bound 58.0% (850/1,466). Screening-detected cases are included; patients with conditions in more than one system are counted in each. N = 1,466 analysed.

## Discussion

A culturally adapted, team-delivered traffic-light lifestyle intervention was feasible in a busy teaching-hospital clinic, with sustained uptake and high responder-reported benefit. This extends the acceptability and clinic-experience findings reported in [16,28] by adding quantified outcomes. The final extract yielded a monotone dose gradient from 5.2% at dose 0 to 98.2% at dose 4 (responder estimate). Among the patients practising all three of the lifestyle advice, revisit patients reported relief more often than single-visit patients (87.2% [395/453] vs 80.8% [455/563]; difference 6.4 percentage points, 95% CI 1.9–10.9). This revisit advantage, like the dose gradient, is consistent with a genuine behaviour–benefit link, but also with selection effects and with more complete ascertainment among patients who return. Proposed common mechanisms (gut motility and barrier function, the brain– gut axis and microbiome, anti-inflammatory and antioxidant effects, metabolic effects, autonomic and neuroendocrine effects, and dietary fibre) remain hypotheses [29,30]. Our intervention components have supportive randomised evidence elsewhere (plant-based diets [7], Yoga for hypertension [8] and diabetes [9], and structured diabetes prevention in Asian Indians [6]); family-adapted programmes improve engagement [31], and self-efficacy mediates adherence [15,32]. These findings suggest that the combined lifestyle programme may help relieve symptoms in patients with chronic diseases, although the single-arm design cannot establish causality.

### Context

Dietary diversity, rather than supplementation alone, is the sustainable route to micronutrient adequacy in India [33]. Score-based, person-centred counselling of this type is typically cost-neutral or cost-saving [34]. The multimorbidity burden and the broadly similar relief pathways observed here are consistent with India’s multimorbidity literature [12].In public-sector health delivery, operationalising dietary interventions requires adapting international Food as Medicine (FAM) constructs to local socio-economic realities. The clinic’s 21-item traffic-light guideline functions as an accessible, low-cost implementation of a whole-food, plant-predominant dietary pattern. Rather than relying on commercial meal delivery, it guides industrial worker families toward affordable, locally available whole plant foods, fresh salads, and culinary spices while eliminating ultra-processed red-category items.

### Comparability with worker populations

This clinic cohort is largely self-selected. For context, cohort prevalence estimates were of similar magnitude to independent regional estimates for industrial workers. A survey of 323 Faridabad factory workers found hypertension in 48.6%, diabetes in 10.5% and obesity in 31.8% [35]. Camp-based screening of 2,588 Faridabad workers found hypertension in 30.9% [36]. The state-wide Haryana STEPS survey found elevated blood pressure in 26.2%, raised blood glucose in 15.5% and overweight or obesity in 35.2%, with tobacco use in 38.9% of men [37]. Corresponding cohort figures, including screening-detected cases, were hypertension 34.9%, diabetes 16.8% and obesity 27.1%. Tobacco abuse (7.8% of the analysed cohort; 16.2% of the 703 men) was below the 41% reported among male industrial workers [38] and the 38.9% among Haryana men [37]. The disease profile in the study cohort is consistent with under-recording of behavioural risk factors in routine care rather than a lower habit burden. Tobacco abuse was nonetheless the most frequent behavioural-risk code in the cohort, and tobacco users who engaged with the programme appeared to respond well: 77 of the 86 tobacco-abuse patients with a codable relief vote (89.5%) reported relief (symptom relief; cessation was not verified).

### Choice environment

Patients visited the IHC exercising free choice among competing options in a tertiary, multi-speciality hospital where consultations, investigations and medicines are free at the point of use, and where dedicated Traditional and Complementary Medicine (AYUSH: Ayurveda, Yoga and naturopathy, unani, siddha and homeopathy) services are also available. The clinic’s main message to prospective patients was explicit: “Come if you want to restore your health, and roll back disease from root causes through self-effort and lifestyle choices of diet, Yoga and spice infusion.” Participation therefore reflects self-selection toward lifestyle care — a consideration relevant to interpreting the compliance and relief patterns reported here.

### The practice of lifestyle medicine

Lifestyle medicine is not a pill, and it cannot be evaluated or delivered through a one-size-fits-all protocol. It requires a deep inquiry into the patient’s way of life to understand not only the root cause and underlying pathology but also one of the harder questions in medicine: why this patient became ill at this particular time. The art of practice of lifestyle medicine includes understanding the patient’s mental attitude and physical infirmities, using the best available evidence and offering a different perspective: a choice between broad-based health and relief from many of the conditions troubling them, on the one hand, and continued deterioration and the development of related conditions if the offending lifestyle continues, on the other. Patients who make this choice were offered a protocol customised to their condition within the 21-item lifestyle guideline and the three core interventions of spice infusion, diet and Yoga. In the IHC clinic Yoga protocol, some asanas were emphasised for particular conditions (for example, Sarvangasana for haemorrhoids). The clinic also emphasised adequate salad intake in the morning and a light, vegetable-based boiled meal in the early evening, sufficiently before sleep. The advice consistently includes complete avoidance of the seven red-category items (foods and addictions), movement matched to the patient’s readiness, and a daily rhythm aligned with the sun’s diurnal cycle. The skills of patient interviewing, non-judgemental listening, root-cause formulation, motivation and continuity across every call and visit are learned competencies, not incidental by-products. We propose to codify this approach and offer it to interested physicians so that it can be practised more widely; the website lifequality.org.in is one attempt by the authors to make this knowledge openly available and encourage wider uptake [39–41].While delivered within a public-sector tertiary teaching hospital framework serving an industrial workforce, the Integrative Health Clinic (IHC) protocol operationalises the standards described by the American College of Lifestyle Medicine (ACLM) and the broader lifestyle-medicine community [42]. By structuring care around the core pillars of lifestyle medicine—specifically integrating whole-food plant-predominant nutrition, structured daily Yoga and pranayama, and circadian alignment— the clinic moves beyond traditional patient education into active therapeutic lifestyle change (ITLC — structured, clinician-guided modification of daily health behaviours as a prescribed, tracked part of care). Furthermore, the integration of family-centred attendance, group peer demonstrations, and a 0–21 tracked lifestyle score illustrates the feasibility of delivering comprehensive lifestyle medicine within a routine outpatient workflow.

### Respect for the patient

The cardinal principle of the IHC clinic has been to keep the patient foremost. This is reflected chiefly in respect for patients’ faith, beliefs, need for attention and dignity, and in treating them as partners in their wellness journey rather than passive recipients of dispensed medicines. This approach helped build trust and mutual connection and improved compliance with lifestyle advice and practices.

### Patient questions and group attendance

During visits and call, the patients often asked many questions about lifestyle and, especially, about the three interventions. The clinic team encouraged questions and took time to answer them, often collectively. Group consultation is now an accepted part of lifestyle-medicine practice. We combined the repeat-visit with new patients, which allowed knowledge, attitudes and practices to spread across successive patient groups and enabled the team to explain lifestyle practices to the group at once.

### Provider time and training

Individualised lifestyle attention is desirable for every patient with a chronic, lifestyle-related condition, but routine clinic time may not permit it. Training providers in the essentials of lifestyle medicine — including its practical nuances, patient engagement, motivation and workflow management to reduce waiting and keep patients engaged — can therefore help make individualised care feasible at scale.

### Access, literacy and motivation

The beneficiary population — ESIC industrial workers and their families — faces steep barriers to clinic follow-up and telephone contact, which should frame the revisit and response figures. Most workers do 12-hour shifts six days a week, and workplace rules generally bar personal calls during shifts and non-emergency hospital visits. Many workers and family members have limited schooling and, where smartphones are available, limited familiarity with their use. Health-seeking behaviour is largely reactive, and the dominant motive for hospital attendance is access to free medicines, rather than lifestyle counselling. It was correspondingly difficult to persuade newly attending patients, in particular, to value and practise lifestyle change alongside medication. These circumstances partly explain the observed participation: only 702 of the 1,737 patients (40.4%) returned for at least one revisit, and 1,466 (84.4%) had a usable telephone contact and were called; 450 analysed patients never gave a definitive answer (30.7%). The revisit and telephone-response rates are therefore best interpreted in the light of access, literacy and motivational constraints rather than as indicators of how patients valued the programme. The constraints described in this paragraph are clinic observations and were not systematically audited.

### Implications

The traffic-light, COM-B and Yoga components should be retained. The next step is prospective, controlled, disease-specific cohorts with predefined primary endpoints (for example, HbA1c, blood pressure or validated symptom scores), defined intervals, capture of harms, intention-to-treat analyses including non-responders, effect sizes and confidence intervals. No claims of cure, pan-disease efficacy or remission without predefined criteria are being made, and patients were advised to continue prescribed treatment. The four-spice culinary infusion remains under study and is not a proven treatment. A further way forward is population-level access to anonymised records: we propose that anonymised data from ESIC’s Dhanwantri portal be made available in CSV form to interested planners, administrators and researchers, to assess the health of worker beneficiaries, to study prevalent conditions (anaemia, occupational disorders, hypertension and diabetes) and to enable data-driven preventive and promotive planning.

### Strengths of the study

The intervention was delivered uniformly through a published 7-step, COM-B-based protocol with a tracked 0–21 lifestyle score, provider-lead, in-clinic demonstration of Yoga and the four-spice infusion, and structured follow-up calls; the protocol and replication resources are publicly available [16,18,39–41]. High compliance with the three practices was largely a product of this delivery and follow-up protocol and plausibly contributed to the reported relief. Medication use was conservative, yet within guideline-concordant categories, with liberal correction of widespread micronutrient deficiency, keeping lifestyle change as the principal addition to standard care. The analysis reports the full 1,737-patient cohort alongside the 1,466 called patients, explicitly compares the two groups, and uses conservative denominators throughout, so responder estimates are not inflated by assumption.

### Limitations

The main limitation is the absence of an untreated or alternative-treatment comparator; natural variation, regression to the mean, concurrent drug changes, placebo effects and selective follow-up may all have contributed to the observed changes. Because the components were delivered jointly, their effects cannot be attributed separately to diet, Yoga, the infusion or medication. Additional limitations include self-reported binary relief (subject to recall and social-desirability bias and open to individual interpretation because the question was generic), classification based on the most recent definitive answer rather than at a fixed time point, 450 analysed patients without a definitive answer (30.7%), a single centre, contamination from free-text diagnoses and the failure to separate contact mode (telephone, WhatsApp or clinic). No validated disease-specific endpoints or effect sizes are presented, and all proportions carry only simple binomial uncertainty. Burden counts include screening-detected cases identified by pragmatic single-cut thresholds rather than clinical adjudication (thresholds and alternative cut-off checks are described in Methods and Table S3). No efficacy or disease-modifying claim is made for the four-spice infusion or for any individual spice. A three-state check of the underlying records (YES / NO / not recorded) showed that for 450 of the 1,466 analysed patients both the practice fields and the relief answer were not recorded; these are the same patients, and no patient had one recorded without the other. The missingness is therefore not at random, no imputation was attempted, and these patients are reported as a separate stratum rather than as confirmed non-adopters (see the note to Supplementary Table S2).

Future research. The four-spice infusion described here is a precisely described, low-cost, self-administered culinary preparation delivered to all patients as one component of a multi-component lifestyle programme. Because diet, Yoga, the infusion and pharmacotherapy were delivered jointly, and because relief was a single generic patient-reported item, this study cannot attribute any outcome to the infusion, and no efficacy or disease-modifying claim is made for it. Its value at this stage is as a reproducible, culturally familiar exposure that can now be investigated directly. Prior work on the constituent spices provides component-level, predominantly metabolic and preclinical evidence only: meta-analyses of cinnamon and fenugreek for glycaemic control [20,21], a placebo-controlled trial of cumin essential oil on glycaemic and inflammatory indices [22], and a preclinical systematic review of coriander and inflammatory mediators [23]. A randomised crossover trial of a polyphenol-rich seven-spice curry (6 g and 12 g) in healthy adults reported acute changes in gut microflora composition [43]. These studies used isolated spices, defined supplemental doses or a cooked curry matrix, and single or acute exposures - not this home-prepared aqueous infusion; no study has evaluated this combination, this preparation or chronic daily use. We therefore propose a staged research agenda: (i) chemical and microbiological characterisation of the infusion, including batch variability, changes in composition across the three-day replenishment cycle, and the safety of that protocol; (ii) a prospectively registered, controlled, component-isolating study with predefined validated endpoints (bowel habit, pain, glycaemic and inflammatory markers) and systematic harms capture, including tolerability and interactions with concomitant medication; and (iii) only thereafter, evaluation of any role in chronic disease management.Furthermore, to overcome the limitation of joint delivery and strengthen causal inference without compromising routine care, prospective series of N-of-1 (single-patient randomised crossover) trials offer a highly rigorous methodological next step. By alternating randomised blocks of specific lifestyle components—such as comparing periods of plant-based diet and Yoga with versus without the four-spice infusion—N-of-1 designs can isolate component-specific effects, control for regression to the mean, and generate aggregated Bayesian population-level evidence. Blinding to a home-prepared infusion is not feasible, so expectancy effects would remain, and an appropriate washout between blocks cannot be specified until the preparation is chemically characterised. This approach directly aligns with the personalised, root-cause paradigm of Lifestyle Medicine as championed by ACLM and the International Board of Lifestyle Medicine (IBLM).

### Behaviour-change trajectory

The practice of Lifestyle change is difficult, for patients and providers alike. Providers must convince patients of the need to change, while avoiding exaggeration of the consequences of not doing so. The trajectory of behaviour change is uncertain, with pitfalls that include lassitude, doubt, peer influence and misinterpretation or misuse of readily available <u>medical</u> information. To counter these threats, the clinic emphasised regular visits (at 2–3-week intervals) and follow-up by telephone, WhatsApp messages and polls. Yet many patients described the challenges of making lifestyle changes and the delicate social choices involved, and some relapsed after initial success. Encouragingly, patients who experience the benefits of change and sustain it for a reasonable period can follow the desired path with less difficulty thereafter.

## Key conclusions

1. **The process of behaviour change in routine care (KAP/COM-B):** This report describes how knowledge, attitude and practice were built in a busy routine setting by a multidisciplinary team trained in all elements. The 21-item guideline clinic-based Yoga demonstration with a video-series link for home practice, and in-clinic demonstration of the four-spice infusion (with a preparation video) supported compliance. The published 7-step protocol [16], delivered by the multidisciplinary team (sequence in Appendix A), addressed all three COM components — capability, opportunity and motivation [17] — and supported high understanding and compliance [15,16,32]. The team observed peer demonstration in action [32]: repeat-visit patients performed Yoga and described their experience before new cases, doctor-led Yoga sessions drew participants, and patients practising in the clinic visibly gained confidence. This is a plausible engagement mechanism rather than a measured effect.
2. **Whole-body pattern:** Reported relief appeared across a range of chronic conditions spanning many systems, pointing to shared mechanisms such as inflammation, gut health, cardiovascular health, neuro-endocrine balance and mental health [29,30].
3. **Joint delivery:** Though diet, exercise and lifestyle changes were delivered together, making their individual contributions unmeasurable, yet their combine effect might have been the principal factor in contributing to well being and relief. In fact, combined diet-and-exercise approaches have the strongest trial support [6,8,9]. Yet, whether the components acted together to produce the observed difference remains a hypothesis.
4. **Family effect:** Patients were encouraged to bring family members and practise the lifestyle changes together. In total, 274 of 1,412 families (19.4%) were represented by more than one member, and 180 patients (10.4%) first attended after another family member [31]. Joint home practice itself was not measured.
5. **Interrelated systems:** Chronic conditions co-occurred (mean 2.24 patient– condition pairs per patient; 63.9% had conditions in two or more systems), and adherence and reported relief were broadly similar across systems. This is consistent with, but does not establish, a whole-body benefit [12].

## Conclusion

This report provides exploratory, hypothesis-generating evidence of feasibility and responder-reported benefit. Prospective controlled evaluation is required before any claim of effect can be made.

## Supporting information

STROBE-Checklist

## Data Availability

Subject to the limitations imposed by the Ethics committee, all data produced in the present study are available upon reasonable request to the authors

## Declarations

### Ethics and consent

The Institutional Ethics Committee, ESIC Medical College & Hospital, Faridabad (EC Reg. No. EC/NEW/INST/2023/HR/0307). The Committee raised no objection to publication on 18 March 2026 (notification 2 March 2026). On 22 October 2026, the Committee conveyed no objection to the proposed analysis and to matching family members by insurance number. All no-objection determinations are subject to strict confidentiality and anonymity, non-disclosure of identifiable information and compliance with applicable institutional policies. Identifiers were removed before analysis, and only anonymised summary statistics are reported.

### De-identification

Analysis inputs were masked before analysis (no names or mobile numbers were included), and insurance numbers in the source extract were used solely for the family-linkage analysis and appear in no output. The tables, figures and text contain only aggregate, non-identifiable data, consistent with the IEC condition of strict confidentiality and anonymity.

### Data availability

The aggregate tables underlying the results are provided as the annexed tables (Tables 1–5) and in a separate supplementary file (Tables S1–S3); the analysis script is available from the corresponding author on reasonable request. The anonymised patient-level dataset is available to bona fide researchers on approved request, subject to the IEC anonymity condition (no public deposit). Protocol/instrument documents: Zenodo doi:10.5281/zenodo.15814357 (2025 guideline) and doi:10.5281/zenodo.24090390 (2026 comprehensive edition); both CC BY 4.0; no patient-level data deposited.

### Consent (participants and publication)

Not applicable — only anonymised summary statistics without identifiable information are reported, and the IEC granted its no-objection on this basis.

### Trial registration

Not applicable — this was a retrospective observational cohort, not a clinical trial, and it was not registered.

### Authors’ contributions (CRediT)

R.S. — conceptualisation, methodology, formal analysis, investigation, project administration, writing (original draft); K.D.C. — supervision, resources, writing (review and editing); J.C.S. — investigation, supervision, writing (review and editing); R.M. — investigation, writing (review and editing). All authors approved the final version and agree to be accountable for all aspects of the work.

AI and analytical assistance. AI tools (DeepSeek and OpenCode) and custom R and Python scripts were used for preliminary data analysis, generation of tables and figures, drafting of the report, editing of the DOCX manuscript, regeneration of figure images, and reference, citation and duplication audits. Their output was reviewed and corrected in depth by the authors to produce the final R script, tables, figures and text. All patient-level data, classifications and interpretations were verified by the authors; AI tools did not generate or alter patient records or make clinical decisions.

## Acknowledgements

We thank Dr. Bhawna Garg and Dr. Pratyaksh Verma, Senior Residents, Department of Medicine, ESIC Medical College, Faridabad for referring cases, and the MBBS interns who rotated through the IHC during May–October 2026 — Abhishek Grewal; Akanksha Verma; Aman; Ankit; Anu; Anuj; Aryan; Ashish Gupta; Avni; Bhawna; Bhumika; Deepanshu Mittal; Deepika; Digdarshini; Dikshit; Evelyn; Garima Yadav; Gautam Yadav; Giriraj; Gopika; Harjeet; Harsh Sharma; Harsh Yadav; Harshit Dahiya; Hemant Kumar; Himanshu; Hitesh; Hrithik; Keshav; Kritika; Kuldeep; Kusum; Liza; Lokesh Attri; Lokesh Garg; Mandeep; Manisha Sharma; Mansi; Mohit Lalwani; Moni Ranga; Monika Yadav; Sushila Mor; Vivek Kumar — for collecting laboratory data, entering case records and contacting patients for follow-up. All of them consented to be acknowledged.

## Administrative basis

The IHC was formalised by a Dean’s office circular (ESICMCHFBD/Dean/Misc/2024) dated 21 December 2024, with a Lifestyle Clinic standard operating procedure (six lifestyle-medicine pillars; multidisciplinary staffing); source documents are on file.

- Reporting guideline: The completed STROBE checklist [26] is provided in a separate file.

## Supplementary material

Aggregate result tables (system-level demographics, dose–relief gradient counts and the screening contribution) are provided in a separate supplementary file (Tables S1–S3); the file contains aggregate counts only — no patient-level data.

## Appendix A Clinic implementation sequence (7-step protocol)

*Based on the published 7-step administration protocol [16] and clinic programme materials*.

1. **Entry and context-setting:** Patients entered through speciality referral or word of mouth. Registration was followed by free consultation, investigation and treatment, with free movement across hospital departments. Lifestyle and nutritional assessment began by asking about the patient’s own views on the role of lifestyle in health [16].
2. **Traffic-light education:** The clinician explained the disease and its lifestyle determinants, then presented the 21 items under red (avoid), green (consume safely) and yellow (lifestyle actions), with open discussion of the evidence, rationale, choices and consequences [16].
3. **Baseline lifestyle score:** The patient graded current practice on the 21 items, and the sum yielded the 0–21 lifestyle score, logged at the first and subsequent visits [16].
4. **Barriers, affirmation and goal-setting:** Barriers and opportunities were explored, and the patient identified changes they were willing to make and set a target score. Lifestyle goals were agreed with the patient (and family) alongside the standard pharmacotherapy plan [16].
5. **Physician reinforcement and demonstration:** The physician reviewed the plan and re-emphasised the lifestyle components. The preparation of the four-spice “power mix” was demonstrated in the clinic with a preparation video, and Yoga asanas and pranayama were demonstrated in the clinic, with a video-series link shared for home practice [16,18,40].
6. **Supervised practice and peer demonstration:** The physician or an on-site trainee or paramedic guided practice with live demonstration and hands-on support. Patients practised exercise or Yoga in the clinic, doctor-led Yoga sessions were held, and repeat-visit patients demonstrated and described their experience before new cases [16].
7. **Follow-up and reinforcement:** Follow-up calls after a few weeks assessed compliance and response. Repeat visits, telephone calls, WhatsApp messaging and mobile polls sustained engagement; the lifestyle score was tracked, and family members were encouraged to join and practise together [16].

