## Supplementary-Tables for "Real-World Outcomes of a Traffic-Light Approach to Lifestyle Modification in Chronic Disease: Evidence from an Integrative Health Clinic"

### Supplementary Tables — Integrated Health Clinic (IHC) Cohort Report

Companion to the revised manuscript `ihc-report-2026-11-05-revised.docx`. Incident cutoff 2026-09-30; analysis extract 2026-11-05. Cohort definitions follow the main report Methods. All tables are aggregate counts; no patient identifiers are included.

#### Table S1. System-level demographics and revisit

| System | Patients | F | M | F age, mean (SD) | M age, mean (SD) | Revisit n | Revisit % |
|---|---|---|---|---|---|---|---|
| ENDOCRINE / METABOLIC | 636 | 394 | 242 | 43.7 (13.0) | 47.9 (12.9) | 338 | 53 |
| CARDIOVASCULAR | 535 | 238 | 297 | 47.5 (12.5) | 50.2 (12.4) | 272 | 51 |
| MUSCULOSKELETAL | 479 | 288 | 191 | 46.7 (12.3) | 48.7 (13.0) | 276 | 58 |
| GASTROINTESTINAL | 340 | 143 | 197 | 39.7 (13.1) | 40.0 (13.5) | 182 | 54 |
| BEHAVIORAL / MENTAL | 167 | 29 | 138 | 42.2 (15.9) | 44.8 (11.9) | 84 | 50 |
| GYNAE / REPRODUCTIVE | 162 | 148 | 14 | 31.6 (10.1) | 36.6 (6.3) | 86 | 53 |
| DIETARY / NUTRITIONAL | 161 | 118 | 43 | 32.5 (15.8) | 29.1 (17.4) | 86 | 53 |
| RESPIRATORY | 118 | 48 | 70 | 39.9 (16.2) | 50.8 (19.3) | 63 | 53 |
| NEUROLOGICAL | 89 | 61 | 28 | 35.6 (13.4) | 48.6 (16.0) | 47 | 53 |
| RENAL / UROLOGY | 81 | 38 | 43 | 38.2 (16.6) | 46.6 (17.4) | 55 | 68 |
| INFECTIONS | 61 | 29 | 32 | 28.4 (16.6) | 38.5 (21.9) | 36 | 59 |
| SKIN | 47 | 19 | 28 | 30.4 (16.3) | 42.2 (14.7) | 21 | 45 |
| NEOPLASTIC / CYSTIC | 15 | 9 | 6 | 41.4 (11.2) | 43.8 (16.7) | 8 | 53 |
| EYE / ENT | 12 | 7 | 5 | 31.6 (22.3) | 32.0 (19.4) | 5 | 42 |

Patients with conditions in multiple systems appear in each system row; F and M counts and revisit n/% are distinct patients with at least one condition in the system. Age means are among distinct patients. Revisit = ≥1 subsequent clinic visit record within the cohort window (call dates are not visits). Screening-detected cases are included; all rows are restricted to the 1,466 analysed patients; rows are ordered by descending patient count.

#### Table S2. Relief by number of lifestyle practices adopted (patient-once)

Practices = spice infusion, diet and Yoga. Per-patient exposure = number of practices coded YES under the most recent definitive answer rule (0–3); the 4-level version additionally assigns 4 when all three practices are YES and the patient has more than one visit (sustained adoption). Relief uses the same rule. Cells with zero patients are shown as 0.

##### (a) 3-level exposure

| Dose (practices) | Relief NO | No definitive answer | YES |
|---|---|---|---|
| 0 | 109 | 450 | 6 |
| 1 | 22 | 0 | 23 |
| 2 | 17 | 0 | 142 |
| 3 | 18 | 0 | 679 |

Dose 0 is a default category, not confirmed non-adoption: 450 of the 565 patients at this dose had no compliance data and were assigned dose 0 because a practice without a YES is counted as not adopted; these patients also have no definitive relief answer.

##### (b) 4-level exposure

| Dose | Relief NO | No definitive answer | YES |
|---|---|---|---|
| 0 | 109 | 450 | 6 |
| 1 | 22 | 0 | 23 |
| 2 | 17 | 0 | 142 |
| 3 (all three practices, single visit) | 12 | 0 | 353 |
| 4 (all three practices, repeated visit) | 6 | 0 | 326 |

Dose 0 is a default category, not confirmed non-adoption: 450 of the 565 patients at this dose had no compliance data and were assigned dose 0 because a practice without a YES is counted as not adopted; these patients also have no definitive relief answer.

All 1,466 analysed patients are represented in each sub-table (3-level: 565/45/159/697; 4-level: 565/45/159/365/332); the 271 patients without a usable telephone contact are compared with the called group in Table 5 of the main report. Figure 2 of the main report plots the responder estimate as YES % (n reporting relief / n with a definitive relief answer at that dose) from sub-table (b); patients with no definitive answer are excluded from this estimate and included in the conservative estimate (dose 0 only: 5.2% vs 1.1%). Dose 0 is a default category, not confirmed non-adoption (see the identical note under each sub-table and the main-report Table 4 footnote). Main-report Table 4 gives both estimates, their denominators, and the dose-0 footnote.

**Note on three-state coding and missing data.** Recoding the underlying records as YES / NO / not recorded for both the practice fields and the relief answer gives the cross-tabulation below (all 1,466 analysed patients; aggregate counts only).

| Exposure record | Outcome ANSWERED | Outcome NOT RECORDED | Total |
|---|---|---|---|
| RECORDED | 1,016 | 0 | 1,016 |
| NOT RECORDED | 0 | 450 | 450 |
| Total | 1,016 | 450 | 1,466 |

The 450 patients with no compliance data are exactly those with no definitive relief answer; no patient has one recorded without the other. Missingness is therefore not at random and no imputation was attempted. As a complete-case sensitivity, excluding these 450 leaves the responder estimates of main-report Table 4 unchanged (5.2 / 51.1 / 89.3 / 96.7 / 98.2 % across dose 0-4) and changes only the dose-0 denominator (565 -> 115) and its conservative estimate (1.1% -> 5.2%). Dose 0 must be read as “no component compliant or compliance not recorded”, not as confirmed non-adoption.

#### Table S3. Screening contribution to main burden counts — leading conditions

| System | Condition | Cases coded | Objective-evidence additions | Cases total |
|---|---|---|---|---|
| ENDOCRINE / METABOLIC | Obesity | 236 | 161 | 397 |
| ENDOCRINE / METABOLIC | Diabetes Mellitus | 244 | 2 | 246 |
| CARDIOVASCULAR | Hypertension | 472 | 39 | 511 |
| DIETARY / NUTRITIONAL | Anaemia | 55 | 44 | 99 |
| **Total** | | **1007** | **246** | **1253** |

"Coded" = patients with the condition recorded as a diagnosis; "Objective-evidence additions" = patients meeting the objective criteria in the main report Methods without a corresponding coded diagnosis at that visit. These pairs are included in the total counts and in main-report Table 2 (see the Table 2 footnote). In total, objective-evidence screening contributed 246 pairs in 233 patients. The Total row sums the four leading-condition rows; entries are condition-level counts (patient-condition pairs), not distinct patients, so a patient with more than one of these conditions is counted in each row.
