## Supplementary material for "Real-World Outcomes of a Traffic-Light Approach to Lifestyle Modification in Chronic Disease: Evidence from an Integrative Health Clinic": STROBE-Checklist

### STROBE Checklist — IHC Faridabad 20-month cohort report

Companion to `ihc-report-2026-11-05-revised.docx` (version dated 5 November 2026). Reporting guideline: von Elm E, et al. STROBE statement (Ann Intern Med 2007;147:573–577) — Ref [26] of the report. Cohort-study version applied; matching/comparator items are not applicable to this single-arm design (noted below). "Location" refers to sections of the 5 November 2026 report.

| # | STROBE item (abridged) | Location | Status / note |
|---|---|---|---|
| 1a | Study design in title/abstract | Title; Abstract/Methods | Yes — "Real-World Evidence"; "Retrospective single-arm cohort". |
| 1b | Informative structured abstract | Abstract | Yes — Background/Methods/Results/Conclusions with key numbers. |
| 2 | Scientific background and rationale | Introduction | Yes — global and India NCD burden [1–3]; US chronic-disease burden and costs [4,5]; diet, Yoga and lifestyle trial evidence [6–11]; guideline and 7-step protocol background [2,16,17]; prior single-disease signals from this clinic [19,24,25]. |
| 3 | Objectives / prespecified hypothesis | Introduction (final paragraph); Methods "Design and aims." | Yes — explicit aim to assess feasibility, compliance with the three lifestyle components and patient-reported relief, and whether the programme could be delivered within routine family-medicine care; restated in "Design and aims.". Exploratory; no prespecified hypothesis. |
| 4 | Study design | Methods "Design and aims." | Yes — retrospective single-arm cohort; STROBE cited [26]; each outcome coded once per patient by the most recent definitive answer rule. |
| 5 | Setting, locations, relevant dates | Introduction; Methods "Setting and dates." | Yes — ESIC Medical College & Hospital, Faridabad; ESI industrial workers and their families; free care at the point of use; 20-month incident period 1 February 2025 to 30 September 2026 with follow-up records through 4 November 2026; incident cut-off (first visit on or before 30 September 2026); October 2026 records contributed follow-up only; January 2025 records were lost in a hospital system upgrade. |
| 6a | Eligibility criteria | Methods "Setting and dates."; "Participants. and diagnosis coding."; Results "Flow."; Table 1 | Yes — a valid record required a physical visit with at least one coded diagnosis field (empty, "NA" and "0" entries excluded); 98 patients (116 records) with a first visit after the cut-off were excluded; 183 records without a coded diagnosis were excluded; all eligible treated patients retained, not only successes; outcome analyses restricted to the 1,466 patients with a usable telephone contact who were called. |
| 6b | Sources and methods of selection | Methods "Setting and dates."; "Participants. and diagnosis coding." | Yes — consecutive electronic clinic record extract; entry by departmental referral or word of mouth; no matching (single arm — not applicable). |

| # | STROBE item (abridged) | Location | Status / note |
|---|---|---|---|
| 6c | Follow-up methods | Methods “Outcomes.”; “Response attribution.”; “Clinic workflow and time per patient.”; Appendix A | Yes — repeat visits, telephone calls and WhatsApp; records ordered by call date; the most recent definitive answer (YES/NO) taken as the patient’s code; calls without a definitive answer (“no response”, “not interested”) were skipped; patients with no definitive answer on any call retained as such, not as NO. |
| 7 | Variables: outcomes, exposures, confounders | Methods “Outcomes.”; “Dose levels.” | Outcomes: patient-level relief (YES/NO/no definitive answer); secondary: patient-level compliance with spice infusion, diet and Yoga. Exposure: five-level dose 0–4 built from the three practices plus a repeat visit. No confounder-adjusted analysis (single-arm design; correlation reported). |
| 8 | Data sources / measurement | Methods “Participants. and diagnosis coding.”; “Screening definitions.”; “Relief measurement.”; Table 1 note | Yes — 699 raw diagnosis labels mapped to 323 canonical conditions in 14 systems; three diagnosis fields; call-date field; BMI, HbA1c, blood-pressure and haemoglobin fields for objective-evidence screening; insurance numbers used only for family linkage and absent from patient-level output; analysis inputs masked (no names or mobile numbers); relief elicited with a single generic question. |
| 9 | Efforts to address bias | Discussion “Limitations.”; “Strengths of the study.”; “Choice environment.”; “Access, literacy and motivation.”; “Behaviour-change trajectory.” | Yes — the full 1,737-patient cohort is reported alongside the 1,466 called patients, with an explicit Table 5 comparison of the two contact groups and conservative denominators throughout; caveats on self-selection, joint delivery, recall/social-desirability bias, single centre, screening thresholds and access barriers are stated. |
| 10 | Study size | Methods “Setting and dates.”; Results “Flow.”; Table 1 | Yes — census of all eligible patients in the incident window (no a-priori sample-size calculation); 4,047 valid records from 1,737 unique patients; 1,466 analysed; 271 patients without a usable telephone contact compared in Table 5. |
| 11 | Quantitative variables | Methods “Dose levels.”; “Screening definitions.”; Table 4; Table S4 | Yes — counts and percentages with explicit denominators; five-level dose 0–4; responder estimate and conservative estimate; pragmatic screening thresholds (BMI >28 kg/m², HbA1c ≥6.5%, ≥2 readings with systolic ≥140 mmHg or diastolic ≥90 mmHg, haemoglobin <10 g/dL) with alternative cut-off checks. |

| # | STROBE item (abridged) | Location | Status / note |
|---|---|---|---|
| 12a | Statistical methods | Methods “Statistics.” | Yes — descriptive only, explicit denominators and column percentages; most recent definitive answer rule; responder estimate (n = 1,016) and conservative lower bound (n = 1,466); 95% CI for the headline proportion; phi correlation. |
| 12b | Subgroup/interaction analyses | Methods “Statistics.” | No subgroup or interaction analyses were performed; disease-specific paired analyses with effect sizes, confidence intervals and sensitivity analyses are deferred to follow-up papers. |
| 12c | Confounder adjustment | Not applicable | No multivariable or adjusted analysis in this exploratory single-arm round; acknowledged in “Statistics.” and “Limitations.”. |
| 12d | Missing data handling | Methods “Outcomes.”; “Statistics.”; Results “Contact, outcomes and adherence.”; Table 3 note; Table 4 note | Yes — 450 analysed patients (30.7%) with no definitive answer retained as such, not imputed; 26 “not interested” retained as no definitive answer, not NO; 271 patients without a usable telephone contact excluded from outcome analyses but retained in cohort flow and Table 5; no best- or worst-case estimate. At dose 0, 450 of 565 patients had no compliance data and defaulted to dose 0, giving the 1.1% (6/565) conservative floor with an explicit footnote. |
| 12e | Loss to follow-up | Methods “Setting and dates.”; Results “Flow.”; Table 1 | Single-visit patients were included without modelled benefit; records after 30 September 2026 are follow-up only; patients whose first visit fell after the cut-off were excluded (98 patients, 116 records); the 271 patients without a usable telephone contact are described in Table 5. No loss-to-follow-up modelling was applied. |
| 13a | Participant flow numbers | Results “Flow.”; Table 1; Figure 1 | Yes — 98 patients (116 records) excluded by the incident-window rule; 183 records excluded for no coded diagnosis; 4,047 valid records from 1,737 unique patients (1,035 single-visit, 702 revisit); 1,466 (84.4%) with a usable telephone contact and called; 271 without. Figure 1 plots 4,044 cleaned records dated on or before 30 September 2026 (1,908 patients). |
| 13b | Reasons for non-participation | Results “Flow.”; Table 1; Table 5 | Yes — 183 blank/undiagnosed records; 271 patients without a usable telephone contact (Table 5); 450 analysed patients with no definitive answer on any call; 26 “not interested” retained as no definitive answer. |
| 13c | Matching criteria | Not applicable | Single-arm; no comparator group. |

| # | STROBE item (abridged) | Location | Status / note |
|---|---|---|---|
| 14a | Descriptive characteristics | Results “Baseline characteristics and burden.”; Table 1 | Yes — 904 female (52.0%; mean age 40.3 ± 14.6 years) and 833 male (48.0%; 44.8 ± 15.1 years); 1,035 single-visit and 702 revisit patients; 2.33 visits per patient; 40.4% revisit; 1,466 (84.4%) with a usable telephone contact and called. |
| 14b | Missing data per variable | Methods “Statistics.”; Table 3 note; Table 4 note | Yes — sex and age available for all 1,737 patients; outcome data only for the 1,466 called; 450 patients (30.7%) with no definitive answer; 450 of 565 dose-0 patients had no compliance data; 271 patients without a usable telephone contact compared in Table 5. |
| 15 | Outcome event numbers | Results “Contact, outcomes and adherence.”; Table 3; Table 4 | Yes — 850/1,016 (83.7%; 95% CI 81.3–85.8) responder relief, 166 NO, 450 with no definitive answer; conservative 58.0% (850/1,466); compliance 783 (77.1%) spice infusion, 846 (83.3%) diet, 825 (81.2%) Yoga among the 1,016 who answered; dose gradient from 5.2% (6/115) at dose 0 to 98.2% (326/332) at dose 4. |
| 16a | Unadjusted estimates and precision | Results “Contact, outcomes and adherence.”; Discussion; Table 3 note | Unadjusted proportions with explicit denominators; Wilson score 95% CI 81.3–85.8 for the headline 83.7%; 6.4-percentage-point revisit advantage (95% CI 1.9–10.9); phi 0.55; all proportions carry only simple binomial uncertainty (“Limitations.”); disease-specific effect sizes and precision deferred to follow-up papers. |
| 16b | Category boundaries | Methods “Dose levels.”; “Screening definitions.”; Table 4 | Yes — five-level dose 0–4 defined (dose 3 = all three components at a single visit; dose 4 = all three with at least one revisit); responder and conservative estimate denominators defined; screening thresholds defined. |
| 16c | Outcome events over time | Not reported by design | Patient-once summary; monthly valid-diagnosis records (91–269) and Figure 1 monthly counts are descriptive only. |
| 17 | Other analyses | Results “Full-trio practitioners.”; “Disease-system adherence and relief.”; Table 5; Figures 3–4; Tables S1–S3 | Yes — full-trio analysis (697 patients; 679 of 850 relief reports), disease-system adherence/relief, Table 5 comparison of the two contact groups, family-linkage and screening cut-off checks; no subgroup hypothesis testing. |
| 18 | Key results vs objectives | Discussion (opening); “Key conclusions.”; “Conclusion.” | Yes — feasibility, dose-graded responder benefit, joint-delivery caution, hypothesis-generating framing; no causal claims. |

| # | STROBE item (abridged) | Location | Status / note |
|---|---|---|---|
| 19 | Limitations | Methods “Harms.”; Discussion “Limitations.”; “Behaviour-change trajectory.” | Yes — no comparator, regression to the mean, joint delivery, self-reported generic relief, most-recent-answer rule, 450 patients (30.7%) with no definitive answer, single centre, pragmatic screening thresholds, no validated disease-specific endpoints, simple binomial uncertainty. No harms were reported in this retrospective extraction (documented events only). |
| 20 | Interpretation | Discussion; “Context.”; “Comparability with worker populations.”; “Strengths of the study.”; “Implications.” | Yes — hypothesis-generating interpretation; consistency with trial/meta-analytic evidence [7–11] and independent regional worker estimates [35–38]; no causal claims; the four-spice infusion is explicitly stated to be unproven. |
| 21 | Generalisability | Discussion “Choice environment.”; “Access, literacy and motivation.”; “Comparability with worker populations.” | Yes — single-centre, largely self-selected ESI worker population in a free-care tertiary multi-speciality setting incl. AYUSH services; access, literacy and motivation barriers described; comparison with independent regional worker prevalence estimates. |
| 22 | Funding | Declarations “Funding and competing interests.” | Yes — none; no author or institution received payment or services from a third party, including any affiliation with or financial benefit from the culinary-spice infusion or its producers. |

Notes: the cohort-study STROBE checklist was used rather than the cross-sectional variant (single-arm cohort with a frozen incident window). Items 6b/13c (matching) and 12c (adjustment) are not applicable to this single-arm design; item 16c is descriptive only. Ethics: Institutional Ethics Committee, ESIC Medical College & Hospital, Faridabad (EC Reg. No. EC/NEW/INST/2023/HR/0307); no-objection 18 March 2026 (see Methods “Ethics and approval.” and Declarations “Ethics and consent.”). Tables 1–5 are annexed to the main report; Tables S1–S3 and this checklist are provided as separate files.
